# Topography and environmental deficiencies are associated with chikungunya virus exposure in urban informal settlements in Brazil

**DOI:** 10.1101/2024.05.02.24306518

**Authors:** Catherine Travis, Hernán Argibay, Maysa Pellizzaro, Daiana Santos de Oliveira, Roberta Santana, Fabiana Almerinda G. Palma, Ricardo Lustosa, Fábio Neves Souza, Yeimi Alexandra Alzate López, Mitermayer Galvão Reis, Albert I. Ko, Peter J. Diggle, Michael Begon, Federico Costa, Max T. Eyre, Hussein Khalil

## Abstract

**Background:** Chikungunya virus (CHIKV) is an arbovirus with a significant global public health burden. Delineating the specific contributions of individual behaviour, household structure and the natural and built environment to CHIKV transmission is important for mitigating risk but challenging in urban informal settlements due to their heterogeneous environments. The aim of this study was to quantify variation in CHIKV seroprevalence between and within four urban communities in a large Brazilian city, and identify the respective contributions of individual, household and environmental factors for seropositivity.

**Methodology/Principal Findings:** A cross-sectional serological survey was conducted in four low-income communities in Salvador, Brazil in 2018 to collect individual, household and CHIKV IgG serology data for 1217 participants. Fine-scale community mapping of high-risk environmental features and remotely sensed environmental data were used to improve characterisation of the microenvironment close to the household. We categorised risk factors into three dimensions - individual, household, and environmental and used binomial mixed-effect models to identify associations with CHIKV seropositivity. CHIKV seroprevalence was 4.8%, 6.1% and 4.3% in three communities and 22.6% in one community which had a distinct topographical profile. The only individual dimension variable associated with seropositivity was male sex (OR 1.67, 95% CI 1.11 - 2.36), but several environmental variables, including living in a house on a steep hillside, at medium to high elevations, and with surface water nearby, were associated with higher seropositivity.

**Conclusions/Significance:** Our findings indicate that CHIKV exposure risk can vary significantly between nearby communities and at fine spatial scales within communities, and is likely to be driven more strongly by the availability of mosquito breeding sites rather than human exposure patterns. They suggest that environmental deficiencies and topography, a proxy for several environmental processes including the degree of urbanisation and flooding risk, may play an important role in driving risk at both of these scales.

## Introduction

Chikungunya virus (CHIKV) is an *Alphavirus* arbovirus that has emerged in the 21st century as a disease of global importance due to its frequent and widespread outbreaks and significant global public health burden [1]. Spread by *Aedes albopictus* and *Aedes aegypti* mosquitoes [2], transmission of CHIKV is particularly intense in tropical urban areas [3] where social, environmental and climatic conditions result in a high availability of breeding sites.

The first CHIKV outbreaks in Brazil occurred in the states of Amapá and Bahia in 2014 and within one year all other states in Brazil had confirmed cases [4]. Since then, over 3.6 million cases have been reported in Brazil [5]. Because of concurrent outbreaks of CHIKV, dengue (DENV), and Zika (ZIKV), health surveillance systems in Brazil are frequently overwhelmed and there is difficulty measuring the true burden of CHIKV [6–7].

Deprived urban communities, or informal settlements, in Northeast Brazil are transmission hotspots for CHIKV and other arboviruses [5] and are characterised by high population density, high temperatures and flooding risk, water shortages, and socioeconomic vulnerability [5]. Structural deficiencies, such as the inadequate provision of drainage and sanitation systems, a lack of regular trash collection and inconsistent water supply, are common. These drive mosquito abundance through standing water in the environment, particularly within discarded containers, and the use of on-site water storage systems.

Previous research in urban areas of Brazil has highlighted the importance of social vulnerability and environmental and infrastructural deficiencies as drivers of CHIKV infection, identifying a range of risk factors for seropositivity that include socioeconomic status (SES), living conditions and the structural quality of houses and nearby pavement [4, 8–10]. These analyses have shown the way in which social, economic and environmental processes overlap to drive CHIKV transmission, but they also demonstrate the inherent challenges in delineating the specific contributions of individual behaviour and mobility, household structure, peridomestic environment and the natural and built environment close to and further away from the household.

Accurate characterisation of the processes that drive arboviral infection in humans requires precise measurement of the environmental features that drive mosquito breeding. This is particularly important in urban informal settings which have complex and heterogeneous rural-urban environments that vary significantly over short distances. Capturing fine-scale variation in environmental features is consequently important for estimating the effect of environmental features, ensuring that environmental features are accounted for correctly when estimating the effect of individual and household factors, and for defining the scale and targeting of vector control within communities. This calls for community-level research that supplements survey variables commonly measured at the household location or municipal level with high-resolution community mapped environmental data that captures the fine scale spatial variation in the natural and built environment [11].

The aim of this study was to quantify variation in CHIKV seroprevalence between and within four urban communities in a large Brazilian city, and identify individual, household and environmental risk factors for seropositivity. We aimed to improve characterisation of the microenvironment close to the household by using fine-scale community mapping of high-risk environmental features and remotely sensed environmental data.

## Methods

### Study Site and Participant Selection

We performed a cross-sectional sero-survey in four communities in the north-western periphery of Salvador; Marechal Rondon (MR), Alto do Cabrito (AC), Nova Constituinte (NC), and Rio Sena (RS) with study areas ranging between 0.07 km^2^ and 0.09 km^2^. These underserved and low-income communities, also known as *favelas* or informal settlements, are characterised by poor living and working conditions and high population density [12]. These communities are not homogeneous areas and have significant variation in socioeconomic status and environmental features over small distances [4, 13].

First, a full census of the study areas was conducted and all individuals ≥5 years of age who slept ≥3 nights per week within the study areas and had lived there for at least 6 months were invited to participate in the study. Enrolled participants or their legal guardians provided written informed consent for blood sample collection and participation in a survey for potential risk factors for arboviral exposure.

### Household Survey

Data was collected between March and October 2018 using a previously standardised questionnaire [13]. The survey items included questions regarding demographics, income, assets, household features, and peri-domestic environmental factors such as presence of open sewers, flooding, trash accumulation, household trash and water storage, mosquito sightings, and if agents from the Center for the Control of Zoonoses (CCZ) who provide health education materials and target vector breeding sites had visited in the past year.

### Serologic Evaluation

During household visits, 10 mL of blood was collected from participants and stored between 2°C–8°C, then sent to the Instituto Gonçalo Moniz, Fundação Oswaldo Cruz laboratory. After aliquoting serum from centrifuged samples, the serums were held at −20°C and tested using IgG ELISA [14].

We interpreted both the CHIKV IgG and IgM ELISA results in ordinance with standard manufacturer instructions. CHIKV IgG absorbance/calibrator levels were negative at <0.8, indeterminate at ≥0.8 to <1.1, and positive at ≥1.1; CHIKV IgM absorbance/calibrator levels were negative at <0.9, indeterminate at ≥0.9 to <1.1, and positive at ≥1.1 [4].

### Environmental variables

#### Mapped variables

Trained field teams systematically surveyed all publicly accessible spaces across the four study areas to collect information on the locations of: i) all trash piles covering an area greater than 0.25m^2^; ii) the open sewer system (including major and minor sewers) mapped as lines. Trash piles and sewers were chosen because of their expected contribution to mosquito breeding risk through rainwater collection in discarded containers and pooling of stagnant water [15]. To create environmental risk variables for use in this analysis, the shortest three-dimensional distance to: i) each trash point and ii) the sewer system was calculated for each household.

#### Remotely sensed variables

Relative elevation was calculated as the difference in elevation above sea level between a given household and the lowest elevation household in that community. Land cover rasters were created from fine-scale WorldView-3 satellite images taken in February 2019 (resolution of 0.3m by 0.3m) using the Semi Automatic Classification plugin in QGIS [16]. Land cover rasters were classified into four categories: impervious surfaces, exposed earth, vegetation, and water sources. To assign a measure of nearby standing water (a proxy for breeding site risk) to each household, a 20m buffer around each household the proportion of the area that was classified as the water land cover type was calculated. A Normalised Difference Vegetation Index (NDVI) layer was also created using the red and near-infrared bands of the World-View 3 satellite images [17] to capture variation in plant density and health which can affect mosquito habitats [18]. Once completed for all communities, the mean NDVI value was generated for a 20m buffer around each household. For all of these remotely-sensed environmental variables, a 20m buffer size was chosen to account for the expected clustering distance of the *Aedes aegypti* mosquito in these highly heterogeneous urban settings [19].

### Statistical analysis

The aim of this study was to identify individual, household and environmental risk factors associated with increased risk of CHIKV seropositivity.

Participant age, education and income were considered as both continuous and ordered categorical variables. In the latter case, we grouped age into ranges of 5–15 years, 16-25, 26-45, 46-65, and ≥66 years to account for the relatively young age sample, and education level into 0-5 years of education, 6-9 years, 10-12 years, and upper level (year 13) or above to correlate with primary, secondary, and upper-level schooling years. Racial categories *preto* (Black) and *pardo* (mixed ethnicities) were combined following the Brazilian Institute of Geography and Statistics (IBGE) race definition [20]. Per capita income was calculated as the total monthly household income in BRL including the value of the government assistance programme bolsa familia, where applicable, divided by the number of people living in the household.

Generalised additive models (GAMs) were used to assess whether the relationship between each continuous explanatory variable and CHIKV seropositivity was linear (Appendix A). As there was evidence against linearity for relative elevation and distance to trash, piecewise linear splines were used with a single knot placed at 17m for relative elevation and 50m for distance to trash.

Variables were grouped into 3 dimensions: 1. Individual variables - sociodemographic factors and exposure-related behaviours such as age, race, income, education, and behaviours that may act as proxies for general skin covering and higher potential vector exposure (e.g. walking barefoot outdoors); 2. Household variables that relate to factors inside of or directly surrounding the home of participants such as housing construction type and the functionality of household features e.g. running water; 3. Environmental variables including garbage collection service, distance to trash locations and sewers, relative elevation, NDVI, and water land cover. A full description of survey variables is provided in Appendix B.

A univariable analysis was performed using a mixed-effects logistic regression with random effects to account for clustering at the household level. Variable selection for the multivariable model was conducted for each dimension separately. Models were fitted for all combinations of variables within each dimension and ranked according to their Akaike Information Criterion (AIC) value. The most parsimonious model was chosen for each dimension, defined as the model with the fewest variables within 2AICc of the lowest value. The variables selected from each dimension were then carried into a final round of variable selection in which the same process was repeated for all of these variables to determine which of them would be included in the final model. Age, sex, and community were considered *a priori* confounders and were included in all models.

We performed the data analysis using R version 3.6.3 [21] and packages tidyverse [22], gmodels [23], and lme4 [24].

### Ethics Considerations

Ethical approval for this study was obtained from the Research Ethics Committee at the Collective Health Institute, Federal University of Bahia, Brazil (permit 041/17 2.245.914.17 2.245.914) and the National Commission for Research Ethics, Brazilian Ministry of Health (CAAE: 68887417.9.0000.5030). All participants involved in the study provided written informed consent before data collection.

## Results

In the four communities, there were 2590 eligible individuals, of which 1318 (50.9%) consented to join the study and provided a blood sample for testing. Of the 1318, 1316 (99.7%) had conclusive serological results with 121 participants (9.19%) testing positive for CHIKV-specific antibodies. CHIKV seroprevalence across the four study areas was 6.1% (23/376) in AC, 4.8% (16/337) in MR, 22.6% (69/305) in NC, and 4.3% (13/298) in RS. Households with seropositive individuals are marked in Figure 1, with the majority of positive households in NC and AC found in or close to the 7.1-15.4m elevation areas. In all of the communities, cases were more commonly found above the lowest relative elevation band.

**Figure 1:**
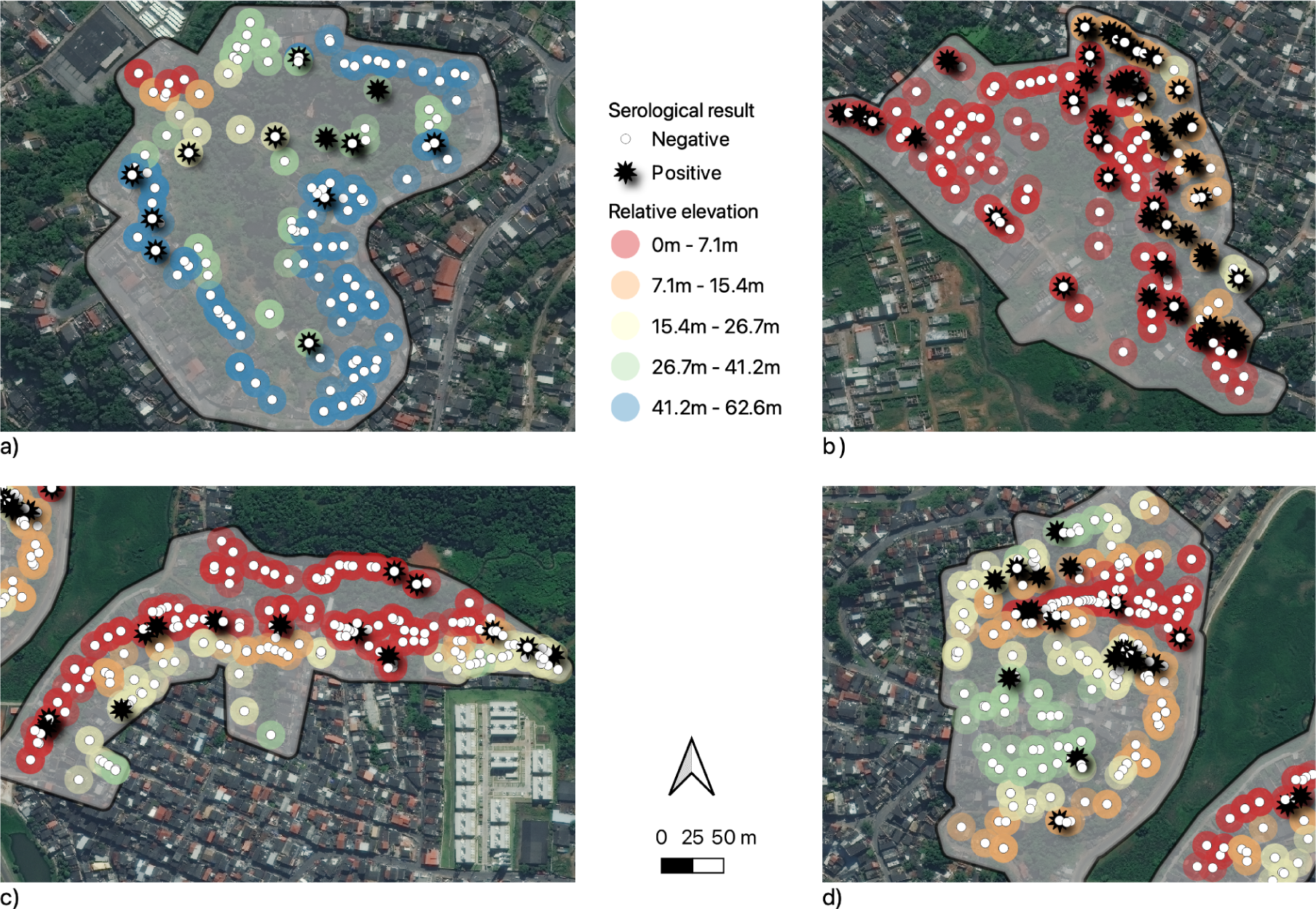
Participant seropositivity (white circle - negative; black star - positive) mapped for each household with household relative elevation shown with a coloured circle for each community: a) Rio Sena, b) Nova Constituinte, c) Marechal Rondon, and d) Alto do Cabrito.

A full description of the study population by each explanatory variable, stratified by serostatus, is provided in Appendix C. Overall, study participants of all areas were relatively young, with median ages ranging from 26 in RS to 38 in MR (IQR 19-48 years). The study population had more women than men (57.3%), was primarily Black (90.88%), and the majority of participants had relatively low income per capita (IQR 12-460 BRL equivalent to 3.3-126.32 USD/month). Three out of the four areas had similar mean absolute household elevations: MR had a mean of 49.62m (IQR 44.0-55.4m) above sea level, AC was 54.74m (IQR 45.0-61.76m), RS was 64.6564m (IQR 57.54-73.00), but NC was far lower than other communities at 8.5473m (IQR 4.83-12.21m).

In the univariable analysis (Table 1), sex was the only individual-level variable that was associated with CHIKV seropositivity, with male participants more likely to test positive for CHIKV than female participants (OR 1.77, 95% CI 1.09 - 2.88). The only variable in the household domain that was associated with seropositivity was whether a household was located on a steep hillside, with participants living in these households having a higher odds of being seropositive (OR 2.76, 95% CI 1.26 - 6.02). Several environmental variables were associated with seropositivity. Participants living in community NC had 10 times the odds of being seropositive relative to those living in MR (OR 10.33, 95% CI 4.24 - 25.22) and participants living in households further away from sewers had a slightly higher odds of being seropositive for each (OR 1.00 per 1m, 95% CI 1.00 - 1.01). The odds of seropositivity increased with increased for each additional metre of household elevation relative to the bottom of each community up to a relative elevation of 17m (elevation 0-17m: OR 1.29, 95% CI 1.15 - 1.44), above which there was no relationship (elevation >17m spline: OR 0.74, 95% CI 0.65 - 0.85; note that this coefficient is relative to the slope estimate for 0-17m).

**Table 1:** Univariable mixed-effects logistic regression model estimates for the probability of a participant being seropositive. Each explanatory variable of interest is grouped by dimension with odds ratios (OR), 95% confidence intervals (95%CI) and p-values shown.

| Variable |  | OR | CI 95% | p value |
| --- | --- | --- | --- | --- |
| Individual Variables |  |  |  |  |
| Sex | Female |  |  |  |
|  | Male | 1.77 | 1.09 - 2.88 | 0.02* |
| Age | 5-15 |  |  |  |
|  | 16-25 | 1.71 | 0.78 - 3.73 | 0.18 |
|  | 26-45 | 1.45 | 0.71 - 2.96 | 0.31 |
|  | 46-65 | 0.87 | 0.37 - 2.04 | 0.75 |
|  | >66 | 1.41 | 0.40 - 4.98 | 0.59 |
| Race | White |  |  |  |
|  | Black/ mixed race | 0.76 | 0.28 - 2.11 | 0.60 |
|  | Asian | 0.83 | 0.11 - 6.17 | 0.86 |
|  | Indigenous | 2.15 | 0.20 - 23.15 | 0.53 |
| Education (years) | 0-5 |  |  |  |
|  | 6-9 | 0.95 | 0.48 - 1.86 | 0.87 |
|  | 10-12 | 0.99 | 0.47 - 2.08 | 0.98 |
|  | Upper Levels (>12) | 0.59 | 0.10 - 3.56 | 0.56 |
| Income per capita (BRL) | 0-99 |  |  |  |
|  | 100-299 | 0.85 | 0.40 - 1.81 | 0.67 |
|  | 300-499 | 0.77 | 0.36 - 1.68 | 0.52 |
|  | 500-1000 | 0.40 | 0.14 - 1.10 | 0.08 . |
|  | >1000 | 0.12 | 0.01 - 1.56 | 0.11 |
| Walk outside barefoot | No |  |  |  |
|  | Yes | 1.42 | 0.80 - 2.52 | 0.23 |
| Household Variables |  |  |  |  |
| House on steep hillside | No |  |  |  |
|  | Yes | 2.76 | 1.26 - 6.02 | 0.01* |
| Lacking water within 30 days | No |  |  |  |
|  | Yes | 0.86 | 0.46 - 1.60 | 0.64 |
| Running water in home | No |  |  |  |
|  | Yes | 1.05 | 0.16 - 6.73 | 0.96 |
| Paved home entry | No |  |  |  |
|  | Yes | 1.64 | 0.78 - 3.43 | 0.19 |
| Wall material | Covered concrete or brick |  |  |  |
|  | Other | 1.17 | 0.46 - 3.00 | 0.74 |
| House with yard | No |  |  |  |
|  | Yes | 0.80 | 0.43 - 1.51 | 0.50 |
| Observed mosquito in home | No |  |  |  |
|  | Yes | 1.14 | 0.61 - 2.14 | 0.68 |
| Water storage in home | No<br>Yes | 1.87 | 0.92 - 3.83 | 0.08 . |
| Water accumulation in home | No<br>Yes | 0.96 | 0.40 - 2.30 | 0.93 |
| CCZ visitation | Within past year<br>Over 1 year /never | 0.50 | 0.23 - 1.08 | 0.08 . |
| Environmental Variables |  |  |  |  |
| Community | Marechal Rondon (MR)<br>Alto do Cabrito (AC)<br>Nova Constituinte (NC)<br>Rio Sena (RS) | 1.31<br>10.33<br>0.90 | 0.54 - 3.18<br>4.24 - 25.22<br>0.33 - 2.42 | 0.55<br><0.001***<br>0.83 |
| Open sewer near house | No<br>Yes | 1.29 | 0.69 - 2.42 | 0.42 |
| Streetlights | No<br>Yes | 0.33 | 0.10 - 1.08 | 0.07 . |
| Water land cover 20m (per 1%) |  | 1.01 | 0.99 - 1.03 | 0.40 |
| Normalised Difference Vegetation Index (per unit) |  | 3.82 | 0.17 - 87.81 | 0.40 |
| Relative elevation (per 1m) | 0-17<br>>17 | 1.29<br>0.74 | 1.15 - 1.44<br>0.65 - 0.85 | <0.001 ***<br><0.001 *** |
| Distance to trash (per 1m) | 0-50<br>>50 | 1.01<br>0.94 | 0.99 - 1.03<br>0.88 - 1.01 | 0.41<br>0.10 |
| Distance to sewer (per 1m) |  | 1.01 | 1.00 - 1.01 | 0.04* |

For the multivariable analysis, 1287 (97%) of the 1316 participants had no missing responses to all variables and were included for this analysis. Each stage of the dimension-specific model selection process is shown in Table 2. Only the three *a priori* confounders (community, sex and age) were maintained for the individual model, water storage in the home, house on a steep hillside, and CCZ visitation were selected from the household model and open sewer, water land cover, and relative elevation were selected for the environmental model. The environmental dimension model provided a better model fit than the individual and household dimension models, as shown by its lower AICc of 706.6. All variables from each of the three dimension models were selected in the final multidimension multivariable model except for water storage in the home and whether there was an open sewer within 10m of the house.

**Table 2:**
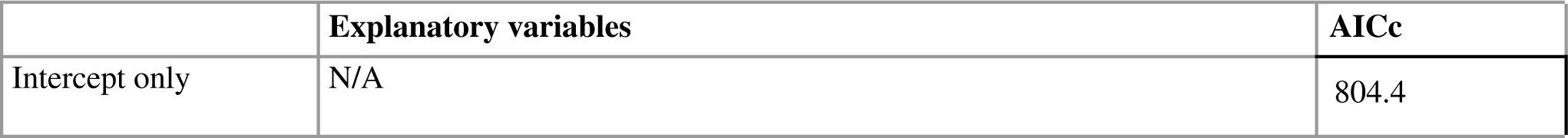
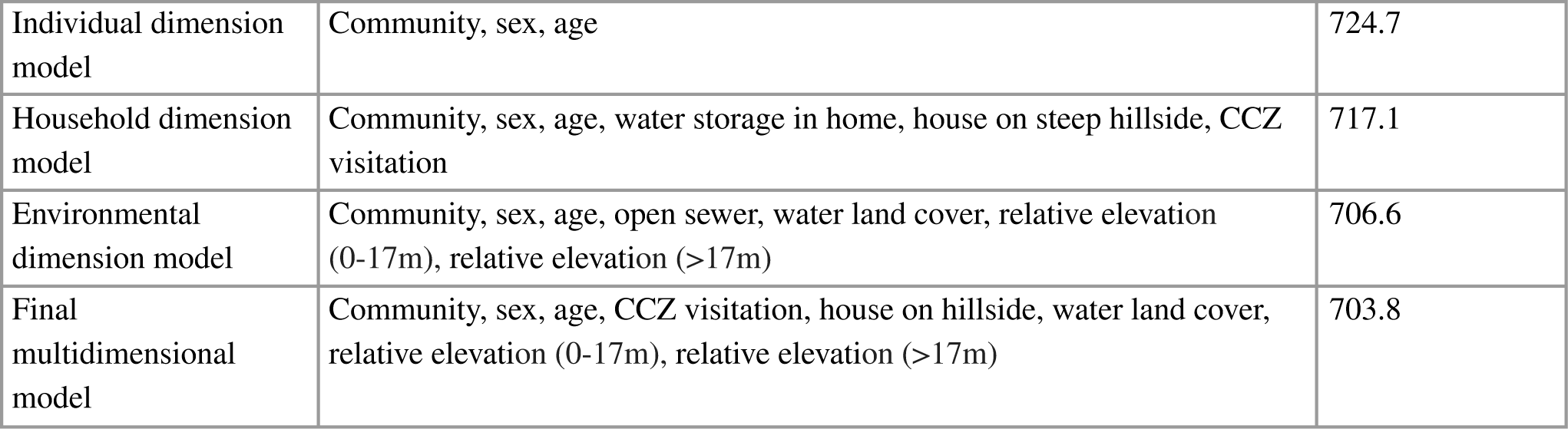
Multivariable model selection by AIC with explanatory variables divided into individual, household, and environmental dimensions for within-dimension variable selection.

|  | Explanatory variables | AICc |
| --- | --- | --- |
| Intercept only | N/A | 804.4 |
| Individual dimension model | Community, sex, age | 724.7 |
| Household dimension model | Community, sex, age, water storage in home, house on steep hillside, CCZ visitation | 717.1 |
| Environmental dimension model | Community, sex, age, open sewer, water land cover, relative elevation (0-17m), relative elevation (>17m) | 706.6 |
| Final multidimensional model | Community, sex, age, CCZ visitation, house on hillside, water land cover, relative elevation (0-17m), relative elevation (>17m) | 703.8 |

In the final multivariable model (Table 3), male participants had a higher estimated odds of being seropositive (OR 1.72, 95% CI 1.12 - 2.65), but there were no clear differences by age group. Participants living in the NC community were much more likely to be seropositive (OR 8.66, 95% CI 4.06 - 18.49 relative to MR community) than participants living in any of the other three communities, all of which had a similar odds of seropositivity. The position of a participant’s household within each community was important for determining exposure risk. Participants living in a house on a steep hillside had an increased odds of seropositivity (OR 2.16, 95% CI 1.15 - 4.03) and those living at the lowest elevation in each community were estimated to have the lowest odds of seropositivity. The odds of seropositivity positively increasing for every additional metre of relative elevation from 0m up to 17m (OR 1.12 per 1m, 95% CI 1.06 - 1.18), above which seropositivity stopped increasing with relative elevation as indicated by a relative odds ratio of 0.84 (OR 0.84 per 1m, 95% CI 0.78 - 0.91) which is equivalent to an absolute odds ratio of 0.98 for each additional metre of elevation above 17m. The proportion of land cover classified as water within a 20m buffer around the household was positively associated with seropositivity (OR 1.01 per 1%, 95% CI 1.00 - 1.02), but CCZ visitation (OR 0.55, 95% CI 0.29 - 1.04) which was selected in the final model was not associated with seropositivity.

**Table 3:** Multivariable mixed-effects logistic regression final model estimates for the probability of a participant being seropositive with odds ratios (OR), 95% confidence intervals (95%CI) and p-values shown.

| Variable |  | OR | CI 95% | p value |
| --- | --- | --- | --- | --- |
| Sex | Female<br>Male | 1.72 | 1.12 - 2.65 | 0.01 * |
| Age | 5-15<br>16-25<br>26-45<br>46-65<br>66+ | 1.84<br>1.48<br>0.99<br>1.49 | 0.93 - 3.64<br>0.79 - 2.77<br>0.47 - 2.08<br>0.50 - 4.44 | 0.08 *<br>0.22<br>0.97<br>0.47 |
| Community | Marechal Rondon (MR)<br>Alto do Cabrito (AC)<br>Nova Constituinte (NC)<br>Rio Sena (RS) | 1.00<br>8.66<br>1.18 | 0.46 - 2.18<br>4.06 - 18.49<br>0.33 - 4.19 | 1<br><0.001 ***<br>0.80 |
| CCZ | Within past year<br>Over 1 year /never | 0.55 | 0.29 - 1.04 | 0.07 . |
| House on steep hillside | No<br>Yes | 2.16 | 1.15 - 4.03 | 0.02 * |
| Water land cover (per 1%) |  | 1.01 | 1.00 - 1.02 | 0.04 * |
| Relative elevation (per 1m) | 0-17m<br>17m+ (relative to 0-17m) | 1.12<br>0.84 | 1.06 - 1.18<br>0.78 - 0.91 | <0.001 ***<br><0.001 *** |

## Discussion

In this multi-community cross-sectional study, we found a low overall CHIKV seroprevalence of 6.1% across the four urban study areas although the community with lowest average elevation, NC, had a significantly higher seroprevalence of 22.6%. We explored individual and contextual (household, environment) risk factors and found that, apart from participant sex, environmental factors such as the elevation and position of a household on a hillside, and the amount of surface water near the household, were most strongly associated with CHIKV seropositivity. Despite our attempts to improve fine-scale measurement of environmental features through the use of high-resolution mapped and remotely sensed variables, only factors related to the environmental context were included in the final model, namely relative elevation and water land cover.

The finding that NC community had a seroprevalence around four times higher than the other three communities presents further evidence that the intensity of CHIKV transmission can vary significantly over relatively small distances between communities and nearby cities. The seroprevalence estimates for the other three communities were lower than the 12% found in a seroprevalence study conducted in other nearby communities in Salvador just over a year earlier (2016-2017) [4], while the high seroprevalence in NC was similar to the 22% found in a recent study (2017-2017) in a nearby city [10]. CHIKV transmission has been characterised by local outbreaks dynamics previously [8, 25], and our findings suggest that NC has suffered a local outbreak, increasing the prevalence of CHIKV-specific antibodies in the population above what was observed in the other areas.

The higher seroprevalence in NC may also be explained by its topography and elevation. The mean elevation above sea level for the communities were 9.8m for MR, 14.7m for AC, 44.6 for RS, and 5.7m for NC. In contrast to the other study areas which are located at higher elevations within steep and narrow valleys, NC is located on a planar surface that is situated at a relatively low elevation. This difference in topography places NC at higher risk of flooding due to the large hydrological catchment area above it, and at a higher risk of standing water forming because of its flat profile limiting water runoff [13, 26]. This is consistent with the finding that more surface water near the household was associated with increased risk of seropositivity. Consequently, while all four study areas have inadequate drainage systems which lead to flooding, NC is likely to be most heavily affected by periods of heavy rainfall and flooding, resulting in a higher availability of short and long-term breeding habitat [27–28] and possibly increased nutrient provision to existing larval containers [29]. Conversely, RS had the highest mean elevation and the lowest prevalence of CHIKV within this study. Identifying communities with these topographical characteristics may be useful for early detection of outbreaks and prioritising areas for vector control in cities.

There was clear evidence that CHIKV exposure risk was also driven by fine-scale variation in the environment within each community. We found that seropositivity was lowest in participants living at the lowest elevation areas of the communities, but that it increased with elevation up to 17 metres of relative elevation. This pattern could also be seen in the maps of household locations with seropositivity marked in Figure 1. In this urban informal setting, living closer to the bottom of the valley is indicative of greater social marginalisation, with households located in these areas found to have lower socioeconomic status [30], poorer WASH infrastructure provision, low-quality housing, inadequate trash disposal and a greater risk of exposure to contaminated floodwater [31]. One explanation for why this may be protective for CHIKV exposure, may be that these low elevation areas are also the least urbanised, consisting of vegetation and soil land cover rather than concrete paving and with a lower population density than higher elevation areas, and are at high-risk of severe and regular flooding with highly contaminated water (due to open sewers and other sources of environmental contamination). This is consistent with the clustering of cases in NC, a high flood-risk community, which were predominantly found at 7-15m relative elevation, above the bottom of the community. This suggests that there may be a relationship with elevation whereby at higher elevations cleaner water is able to pool in plastic containers, trash and paved areas and at lowest elevations there are fewer possible breeding sites due to lower levels of urbanisation and the flooding frequency being so high, or flood water so contaminated with organic material, that it flushes out or contaminates viable breeding sites.

The finding that households located on steep hillsides had a higher risk of seropositivity is consistent with this relative elevation hypothesis. In rapidly urbanising areas like Salvador, informal housing for the most marginalised populations is common on steep hillsides that are not typically considered suitable for building. Households in these areas are located at higher elevations in highly urbanised settings where they are protected from the most intense flooding, but they are generally of very low socioeconomic status and are poorly served by basic urban services [32].

The fact that people living in the lower elevation areas are the most socioeconomically precarious and have been found to have the highest rate of migration in and out of our study communities may also have contributed to the association found with elevation. These residents may have moved from lower prevalence rural areas in the interior of the state of Bahia more recently and had a lower exposure time in the study community. However, a significant proportion of the migration is usually from similar urban areas in Salvador and we would only expect this to contribute to a relatively small degree.

Consistent with previous research of CHIKV in Salvador, we found a significant relationship between male sex and CHIKV seropositivity. This may be a result of men travelling longer average distances within the community during the day than women, as was found in a previous study in Salvador [33], which would give them greater exposure to mosquitoes away from the household than women. The finding that households who had been visited by CCZ more recently had a higher odds of seropositivity suggests that they are identifying and targeting the highest-risk households.

Environmentally transmitted diseases, such as leptospirosis, that have been studied in these areas extensively depend on both individual and contextual risk factors [13] including resident knowledge, attitudes, and practices [34]. However, for mosquito-borne pathogens, transmission depends more on environmental factors promoting vector breeding and presence rather than e.g. individual behaviours in low-income urban settings. Our results show that there are significant risk gradients for CHIKV exposure between and within communities and which is likely to be driven by the availability of mosquito breeding sites rather than human exposure patterns. This availability appears to be driven by topography and environmental deficiencies, including flooding risk, infrastructure provision and the type of environment close to the household, all of which are challenging to measure at high spatial resolutions. While elevation may act as a proxy for these environmental processes in this urban setting, future studies should aim to improve on the suite of high-resolution environmental variables used in this study. In particular, our results show that capturing long-term water bodies is insufficient and there is a need for measuring flooding risk more directly through measures such as the topographic wetness index (TWI) or through identification of short-term breeding habitat using higher temporal resolution satellite imaging after heavy rainfall.

A limitation of this study is its cross-sectional study design. As CHIKV-specific IgG antibodies can remain for over 12 months [35], exposure to infected mosquitoes could have taken place at a time when the environment, household, or individual risk factors we measured may not be fully representative of the potential exposure period. Future longitudinal studies are therefore needed to measure CHIKV incidence and identify fine scale risk factors and gradients at the time of exposure, and will also enable the interplay between meteorological factors, flooding, the environment and CHIKV transmission to be studied at high temporal and spatial resolutions.

This study has shown the importance of identifying environmental risk factors for CHIKV transmission at both community and within-community scales, and highlighted the challenges associated with accurately measuring these environmental processes in highly heterogeneous urban informal settlements. These findings demonstrate how increasingly urbanised and marginalised populations globally are forced to live in structurally and topographically unsafe environments that place them at high risk of arboviral infection.

## Data Availability

All data produced in the present study are available upon reasonable request to the authors.

## Supporting Information Captions

## Appendices

## Appendix A: GAM Graphs

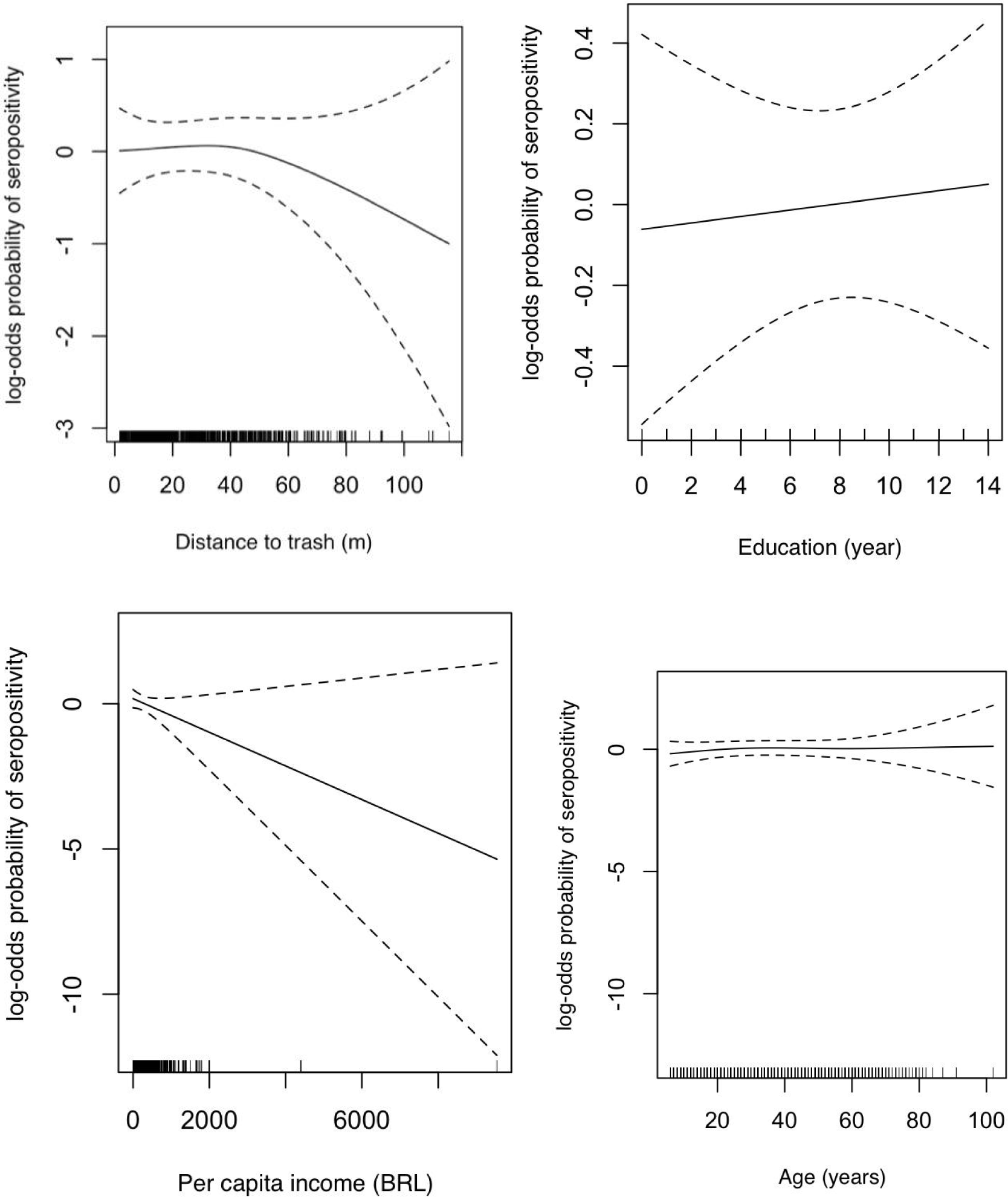
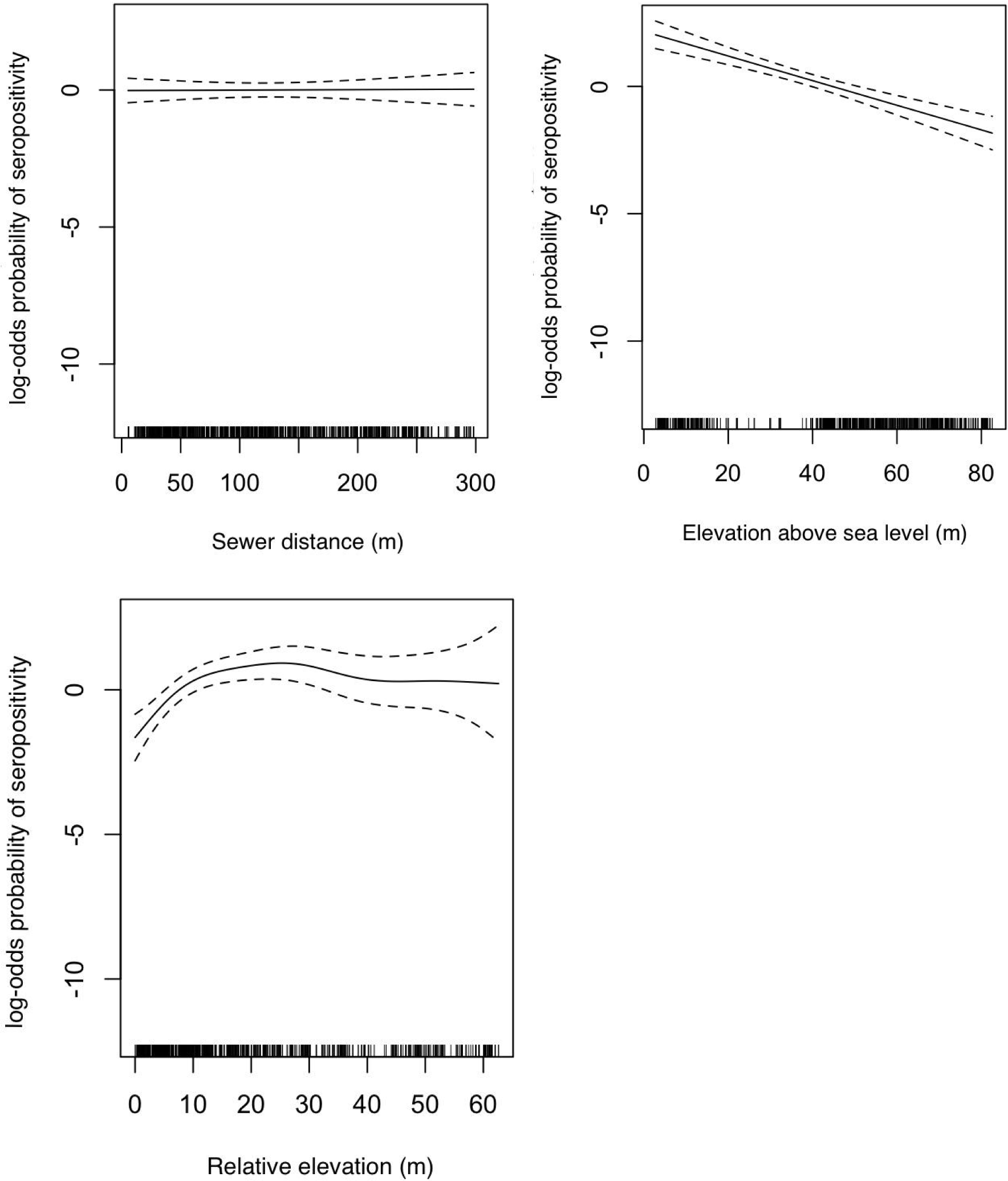

## Appendix B: Variable Descriptions

Responses to these variables are based on information provided from interviews the field team asked the head of each surveyed household. All questions regarding environmental factors were referring to a distance of 10 metres from the house.

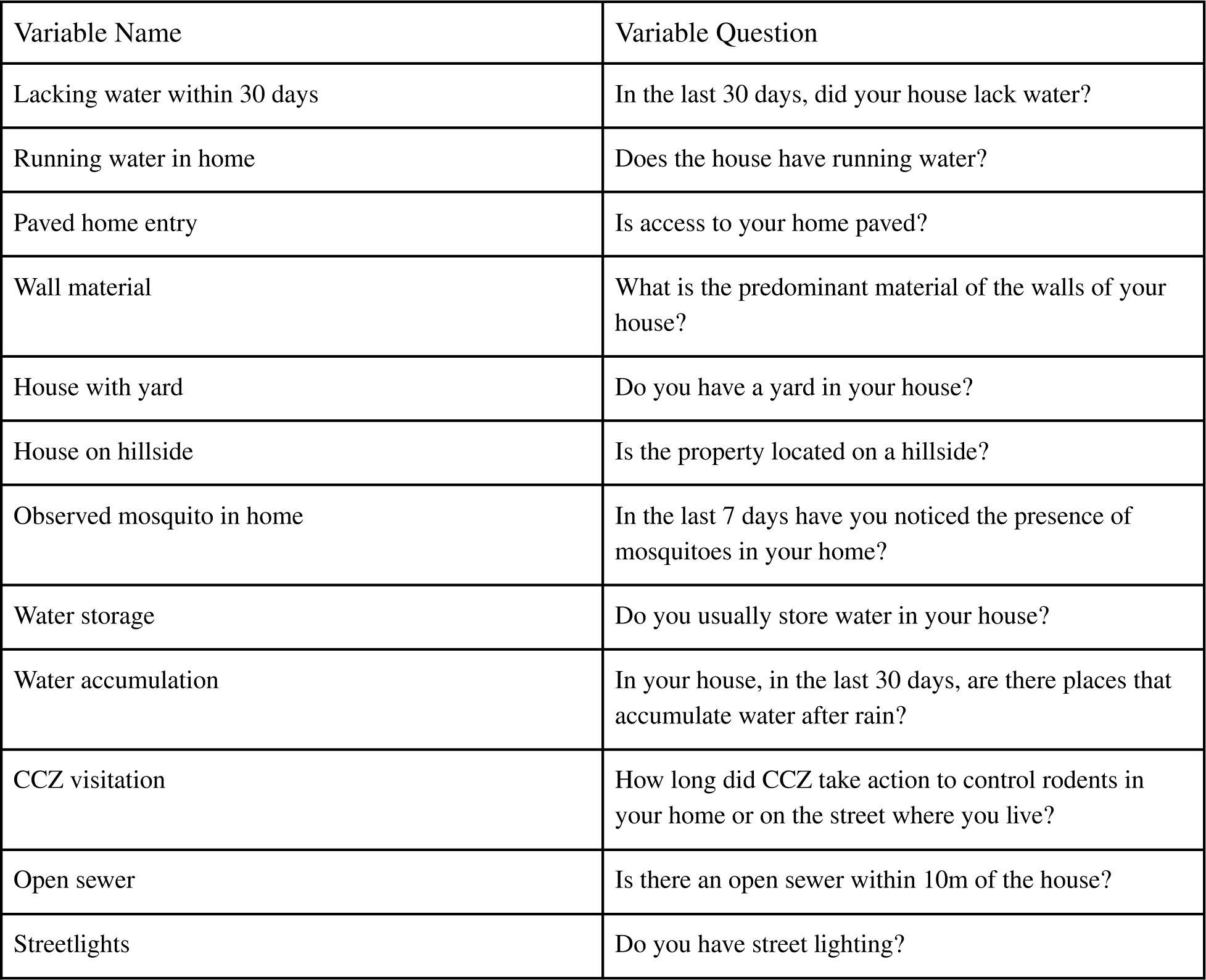

| Variable Name | Variable Question |
| --- | --- |
| Lacking water within 30 days | In the last 30 days, did your house lack water? |
| Running water in home | Does the house have running water? |
| Paved home entry | Is access to your home paved? |
| Wall material | What is the predominant material of the walls of your house? |
| House with yard | Do you have a yard in your house? |
| House on hillside | Is the property located on a hillside? |
| Observed mosquito in home | In the last 7 days have you noticed the presence of mosquitoes in your home? |
| Water storage | Do you usually store water in your house? |
| Water accumulation | In your house, in the last 30 days, are there places that accumulate water after rain? |
| CCZ visitation | How long did CCZ take action to control rodents in your home or on the street where you live? |
| Open sewer | Is there an open sewer within 10m of the house? |
| Streetlights | Do you have street lighting? |

## Appendix C: Variable Counts and Percentages

CHIKV seroprevalence, determined by detection of IgG, stratified by demographic, behavioural, and environmental factors (n=1316).

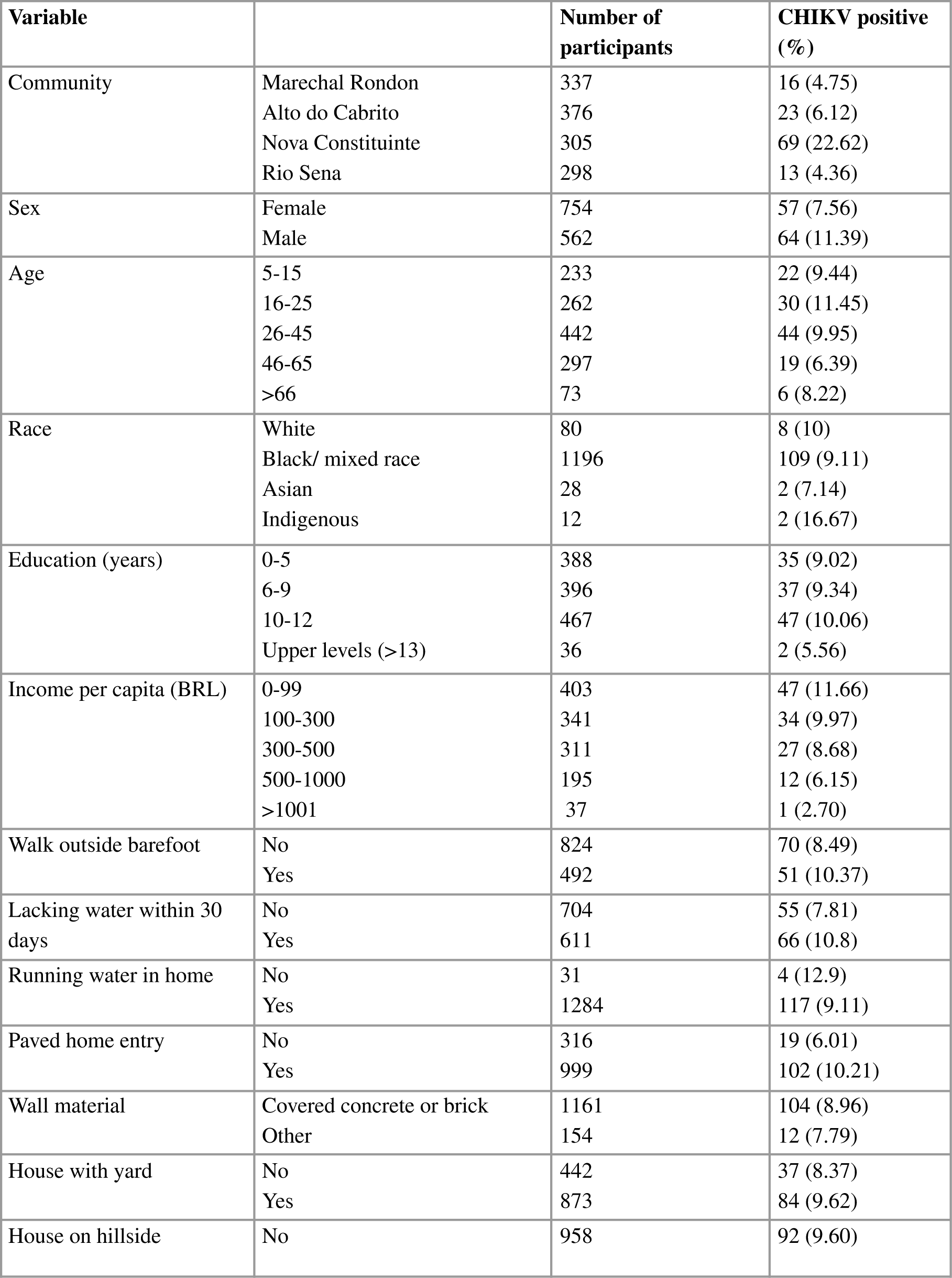
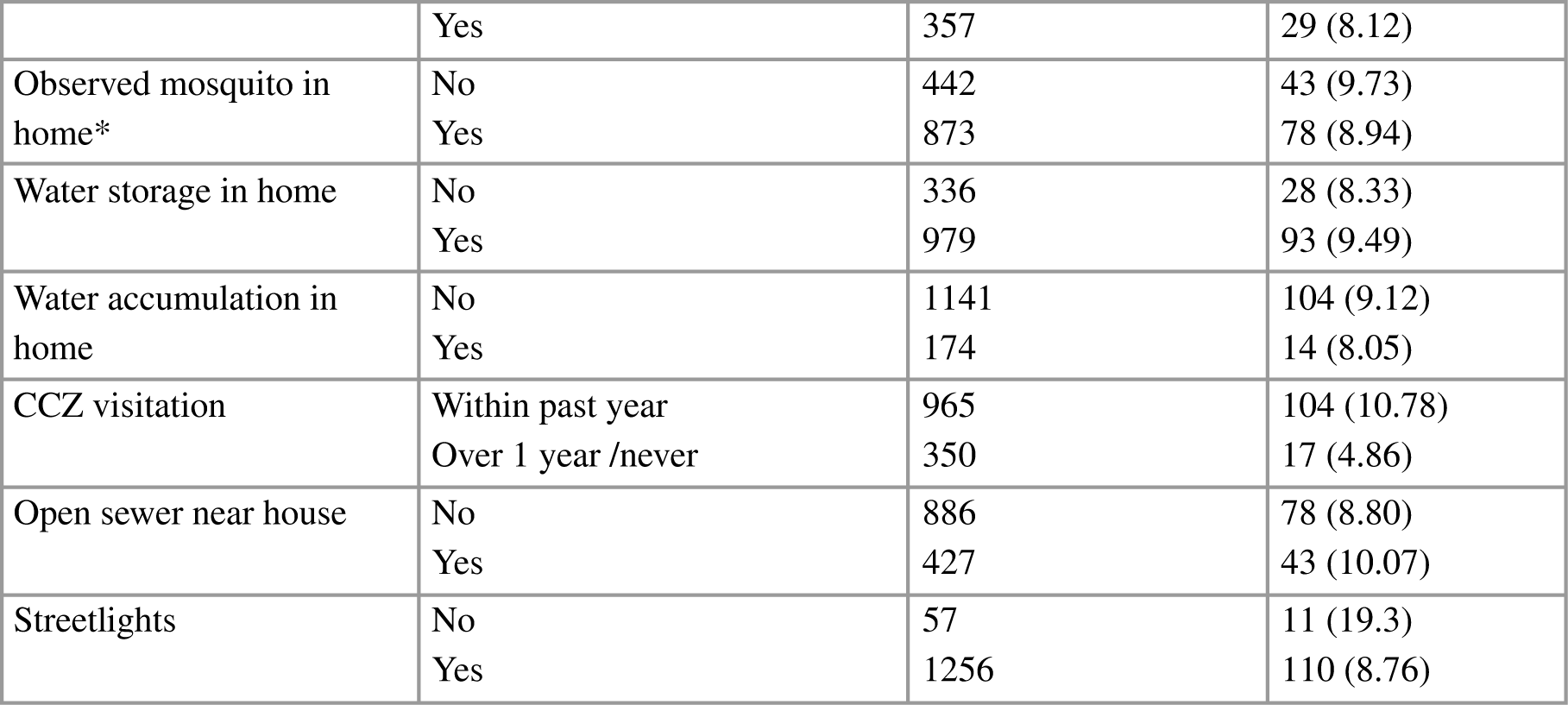

## Appendix D: Model Selection

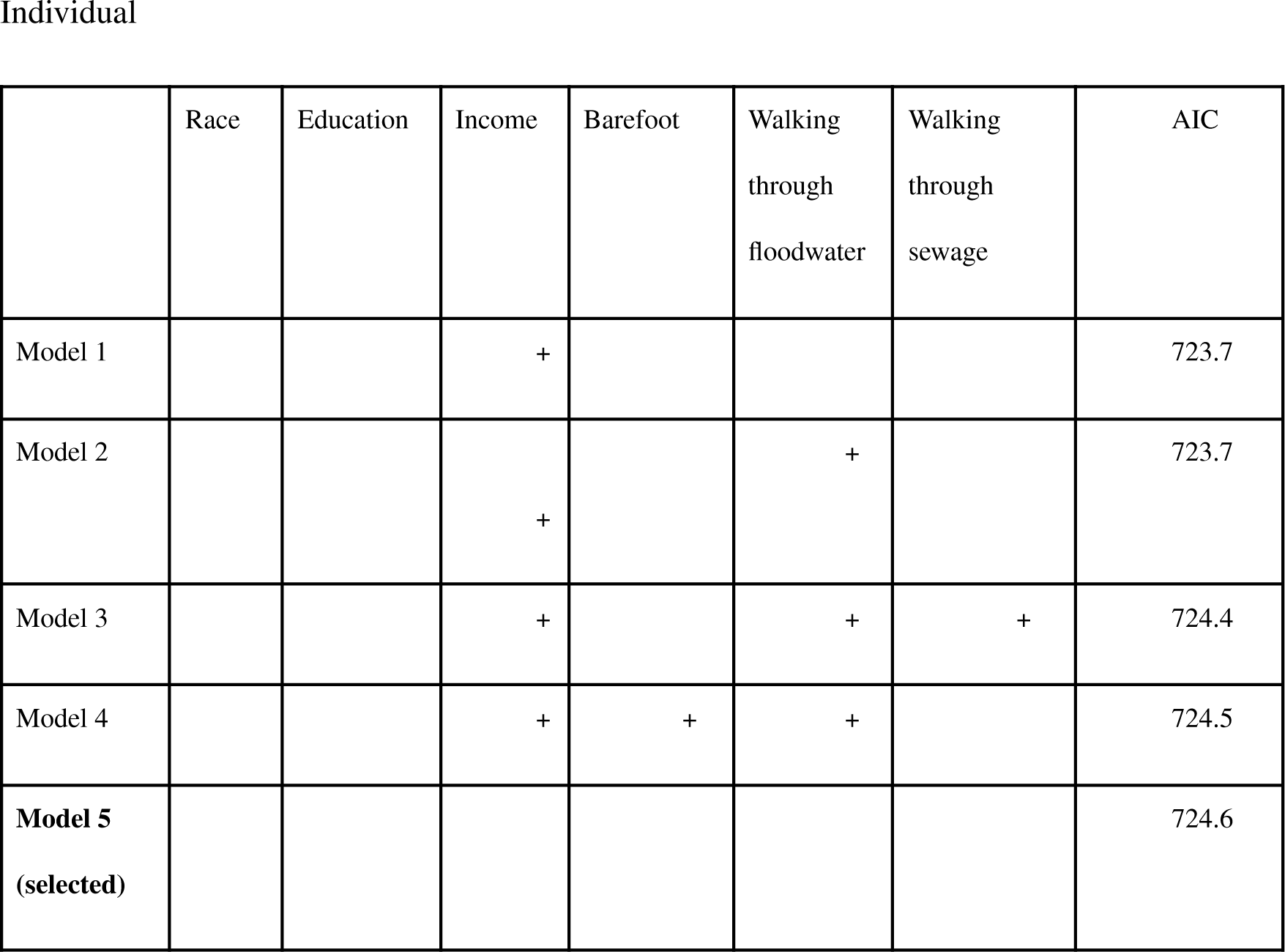
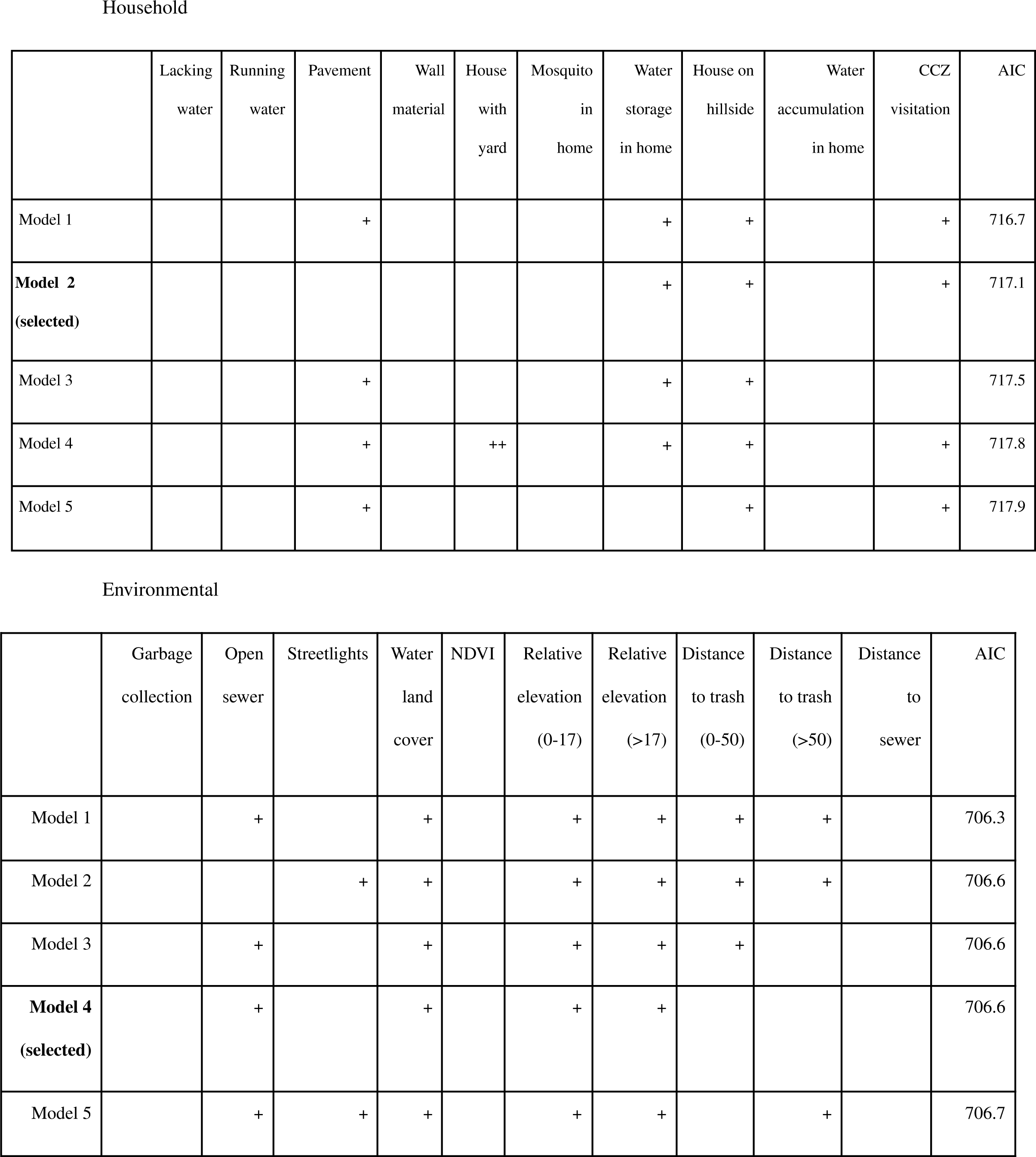
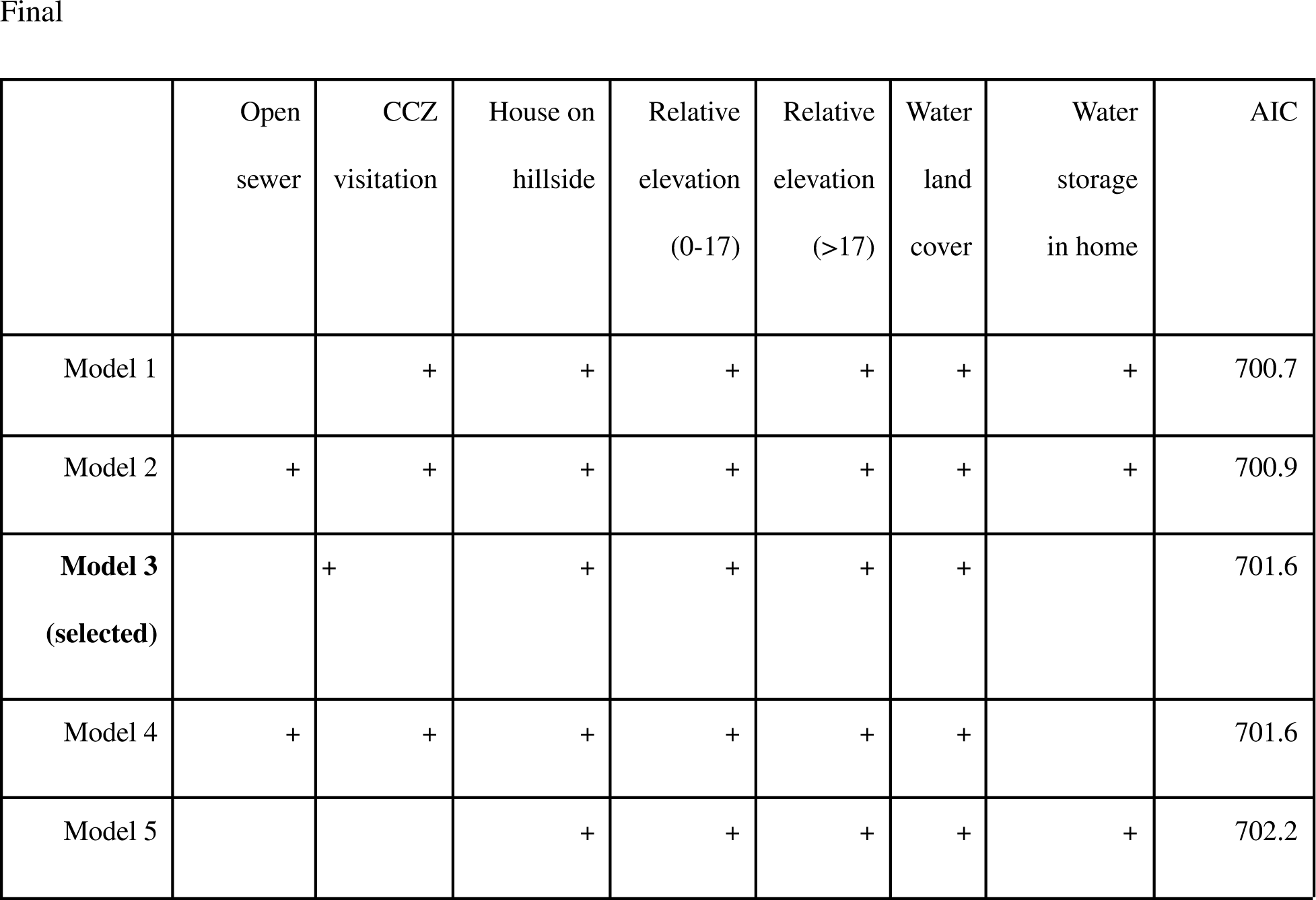

## References

1. Wahid, B., Ali, A., Rafique, S., & Idrees, M. (2017). Global expansion of chikungunya virus: mapping the 64-year history. International journal of infectious diseases : IJID : official publication of the International Society for Infectious Diseases, 58, 69–76. 10.1016/j.ijid.2017.03.006

2. Paupy, C., Ollomo, B., Kamgang, B., Moutailler, S., Rousset, D., Demanou, M., Hervé, J. P., Leroy, E., & Simard, F. (2010). Comparative role of Aedes albopictus and Aedes aegypti in the emergence of Dengue and Chikungunya in central Africa. Vector borne and zoonotic diseases (Larchmont, N.Y.), 10(3), 259–266. 10.1089/vbz.2009.0005

3. Petersen, L. R., & Powers, A. M. (2016). Chikungunya: epidemiology. F1000Research, 5, F1000 Faculty Rev-82. 10.12688/f1000research.7171.1

4. Anjos, R. O., Mugabe, V. A., Moreira, P. S., Carvalho, C. X., Portilho, M. M., Khouri, R., . . . Ribeiro, G. S. (2020). Transmission of Chikungunya Virus in an Urban Slum, Brazil. Emerging Infectious Diseases, 26(7), 1364–1373.

5. Ferreira de Almeida, I., Codeço, C. T., Lana, R. M., Bastos, L. S., de Souza Oliveira, S., Andreza da Cruz Ferreira, D., Godinho, V. B., Souza Riback, T. I., Cruz, O. G., & Coelho, F. C. (2023). The expansion of Chikungunya in Brazil. The Lancet Regional Health - Americas, 25, 100571. 10.1016/j.lana.2023.100571

6. Cardoso, C. W., Kikuti, M., Prates, A. P., Paploski, I. A., Tauro, L. B., Silva, M. M., Santana, P., Rego, M. F., Reis, M. G., Kitron, U., & Ribeiro, G. S. (2017). Unrecognized Emergence of Chikungunya Virus during a Zika Virus Outbreak in Salvador, Brazil. PLoS neglected tropical diseases, 11(1), e0005334. 10.1371/journal.pntd.0005334

7. Neto, A. S., Sousa, G. S., Nascimento, O. J., & Castro, M. C. (2019). Chikungunya- attributable deaths: A neglected outcome of a neglected disease. PLOS Neglected Tropical Diseases, 13(9).

8. Tauro, L. B., Cardoso, C. W., Souza, R. L., Nascimento, L. C., Santos, D., Campos, G. S., Sardi, S., Reis, O., Reis, M. G., Kitron, U., & Ribeiro, G. S. (2019). A localized outbreak of Chikungunya virus in Salvador, Bahia, Brazil. Memorias do Instituto Oswaldo Cruz, 114, e180597. 10.1590/0074-02760180597

9. Dalvi, A. P., Gibson, G., Ramos, A. N., Bloch, K. V., Sousa, G. dos, Silva, T. L., Braga, J. U., Castro, M. C., & Werneck, G. L. (2023). Sociodemographic and environmental factors associated with Dengue, Zika, and chikungunya among adolescents from two Brazilian capitals. PLOS Neglected Tropical Diseases, 17(3). 10.1371/journal.pntd.0011197

10. Teixeira, M. G., Skalinski, L. M., Paixão, E. S., Costa, M. da, Barreto, F. R., Campos, G. S., Sardi, S. I., Carvalho, R. H., Natividade, M., Itaparica, M., Dias, J. P., Trindade, S. C., Teixeira, B. P., Morato, V., Santana, E. B., Goes, C. B., Silva, N. S., Santos, C. A., Rodrigues, L. C., & Whitworth, J. (2021). Seroprevalence of chikungunya virus and living conditions in Feira de Santana, Bahia-Brazil. PLOS Neglected Tropical Diseases, 15(4). 10.1371/journal.pntd.0009289

11. Lowe, R., Lee, S. A., O’Reilly, K. M., Brady, O. J., Bastos, L., Carrasco-Escobar, G., de Castro Catão, R., Colón-González, F. J., Barcellos, C., Carvalho, M. S., Blangiardo, M., Rue, H., & Gasparrini, A. (2021). Combined effects of hydrometeorological hazards and urbanisation on dengue risk in Brazil: A Spatiotemporal Modelling Study. The Lancet Planetary Health, 5(4). 10.1016/s2542-5196(20)30292-8

12. Wallenfeldt, J. (2023, February 16). favela. Encyclopedia Britannica. https://www.britannica.com/topic/favela

13. Khalil H, Santana R, de Oliveira D, Palma F, Lustosa R, Eyre MT, et al. (2021) Poverty, sanitation, and Leptospira transmission pathways in residents from four Brazilian slums. PLoS Negl Trop Dis 15(3): e0009256. 10.1371/journal.pntd.0009256

14. Euroimmun, https://www.euroimmun.com

15. Montalvo, T., Higueros, A., Valsecchi, A., Realp, E., Vila, C., Ortiz, A., Peracho, V., & Figuerola, J. (2022). Effectiveness of the Modification of Sewers to Reduce the Reproduction of Culex pipiens and Aedes albopictus in Barcelona, Spain. Pathogens (Basel, Switzerland), 11(4), 423. 10.3390/pathogens11040423

16. QGIS https://www.qgis.org/en/site/forusers/download.html

17. Hansen, M. C., & Loveland, T. R. (2012). A Review of Large Area Monitoring of Land Cover Change Using Landsat Data. Remote Sensing of Environment, 122, 66–74. 10.1016/j.rse.2011.08.024

18. Weaver, S. C., & Reisen, W. K. (2010). Present and future arboviral threats. Antiviral research, 85(2), 328–345. 10.1016/j.antiviral.2009.10.008

19. Schafrick, N. H., Milbrath, M. O., Berrocal, V. J., Wilson, M. L., & Eisenberg, J. N. S. (2013). Spatial Clustering of Aedes aegypti Related to Breeding Container Characteristics in Coastal Ecuador: Implications for Dengue Control. The American Society of Tropical Medicine and Hygiene, 89(4), 758–765.

20. Osorio, R. G. (2003). O Sistema Classificatório de “Cor ou raça - ipea. Institute for Applied Economic Research. https://www.ipea.gov.br/portal/images/stories/PDFs/TDs/td_0996.pdf

21. https://www.R-project.org/

22. https://www.tidyverse.org/packages/

23. https://cran.r-project.org/web/packages/gmodels/index.html

24. https://cran.r-project.org/web/packages/lme4/index.html

25. Anjos, R. O., Portilho, M. M., Jacob-Nascimento, L. C., Carvalho, C. X., Moreira, P. S., Sacramento, G. A., Nery Junior, N. R., de Oliveira, D., Cruz, J. S., Cardoso, C. W., Argibay, H. D., Plante, K. S., Plante, J. A., Weaver, S. C., Kitron, U. D., Reis, M. G., Ko, A. I., Costa, F., & Ribeiro, G. S. (2023). Dynamics of chikungunya virus transmission in the first year after its introduction in Brazil: A cohort study in an Urban Community. PLOS Neglected Tropical Diseases, 17(12). 10.1371/journal.pntd.0011863

26. Lopo, A. B., Charles Matheus Pinheiro Da Cruz Machado, & Amorim, J. M. (2018). QUALIDADE DAS ÁGUAS DO RIOS URBANOS DE SALVADOR: O caso do Rio das Pedras. Anais Do(a) Anais Da Mostra De Pesquisa Em Ciência E Tecnologia.

27. Ahern M, Kovats RS, Wilkinson P, Few R, Matthies F (2005) Global health impacts of floods: epidemiologic evidence. Epidemiologic Reviews 27:36–46

28. Pires DA, Gleiser RM (2010) Mosquito fauna inhabiting water bodies in the urban environment of Córdoba city, Argentina, following a St. Louis encephalitis outbreak. Journal of Vector Ecology 35:401–409.

29. Yee, S. H., Yee, D. A., de Jesus Crespo, R., Oczkowski, A., Bai, F., & Friedman, S. (2019). Linking water quality to aedes aegypti and Zika in flood-prone neighborhoods. EcoHealth, 16(2), 191–209. 10.1007/s10393-019-01406-6

30. Reis, R. B., Ribeiro, G. S., Felzemburgh, R. D., Santana, F. S., Mohr, S., Melendez, A. X., Queiroz, A., Santos, A. C., Ravines, R. R., Tassinari, W. S., Carvalho, M. S., Reis, M. G., & Ko, A. I. (2008). Impact of environment and social gradient on Leptospira infection in urban slums. PLoS Neglected Tropical Diseases, 2(4). 10.1371/journal.pntd.0000228

31. Eyre, M. T., Souza, F. N., Carvalho-Pereira, T. S., Nery, N., de Oliveira, D., Cruz, J. S., Sacramento, G. A., Khalil, H., Wunder, E. A., Hacker, K. P., Hagan, J. E., Childs, J. E., Reis, M. G., Begon, M., Diggle, P. J., Ko, A. I., Giorgi, E., & Costa, F. (2022). Linking rattiness, geography and environmental degradation to spillover leptospira infections in marginalised urban settings: An eco-epidemiological community-based cohort study in Brazil. eLife, 11. 10.7554/elife.73120

32. Snyder, R. E., Jaimes, G., Riley, L. W., Faerstein, E., & Corburn, J. (2013). A Comparison of Social and Spatial Determinants of Health Between Formal and Informal Settlements in a Large Metropolitan Setting in Brazil. Journal of Urban Health, 91(3), 432–445.

33. Owers KA, Odetunde J, de Matos RB, Sacramento G, Carvalho M, Nery N Jr, et al. (2018) Fine-scale GPS tracking to quantify human movement patterns and exposure to leptospires in the urban slum environment. PLoS Negl Trop Dis 12(8): e0006752. 10.1371/journal.pntd.0006752

34. Palma FAG, Costa F, Lustosa R, Mogaji HO, de Oliveira DS, et al. (2022) Why is leptospirosis hard to avoid for the impoverished? Deconstructing leptospirosis transmission risk and the drivers of knowledge, attitudes, and practices in a disadvantaged community in Salvador, Brazil. PLOS Global Public Health 2(12): e0000408. 10.1371/journal.pgph.0000408

35. Pierro, A., Rossini, G., Gaibani, P., Finarelli, A. C., Moro, M. L., Landini, M. P., & Sambri, V. (2015). Persistence of anti-chikungunya virus-specific antibodies in a cohort of patients followed from the acute phase of infection after the 2007 outbreak in Italy. New microbes and new infections, 7, 23–25. 10.1016/j.nmni.2015.04.002

